# ‘Kindness By Post’: A Mixed-Methods Evaluation of A Participatory Public Mental Health Project

**DOI:** 10.1101/2021.11.19.21266589

**Authors:** Congxiyu Wang, Eiluned Pearce, Rebecca Jones, Brynmor Lloyd-Evans

**Affiliations:** Department of Psychiatry, University of Oxford, Warneford Hospital, Warneford Ln, Headington, Oxford OX3 7JX, United Kingdom; Division of Psychiatry, Faculty of Brain Sciences, University College London, Maple House, 149 Tottenham Court Rd, London W1T 7BN, United Kingdom

**Keywords:** Kindness, Wellbeing, Loneliness, Belongingness, Public Health

## Abstract

**Background:** Random acts of kindness can improve wellbeing. However, less is known about the impacts of giving and receiving acts of kindness with strangers on wellbeing and loneliness. Therefore, this study’s objectives were to evaluate a participatory public mental health project involving sending and receiving a card with goodwill messages, to understand how such acts of kindness influence wellbeing and loneliness, and to investigate the potential mechanisms underlying the project’s impacts.

**Materials and methods:** This study was a secondary analysis of anonymised service evaluation data collected in the ‘Kindness by Post’ (KBP) project in 2020. It used a mixed-methods single-group design and data from 289 participants. Changes in wellbeing, loneliness, sense of belonging and hope from baseline to follow-up were analysed using linear or multinomial logistic regression. Regression models also examined the associations between changes in wellbeing and baseline loneliness or participation level. Free text responses about experiences and suggestions for the project were analysed using thematic analysis.

**Results:** Participants had a small, but statistically significant improvement, in wellbeing equating to 0.21 standard deviations (SD) (95% CI: 0.12 to 0.30) after taking part in the project, as well as improvements in loneliness, sense of belonging and hope. How lonely a participant was at baseline and whether participants both sent and received a kindness card were not associated with improvements in wellbeing. In the qualitative analysis, a desire to help others emerged as the main motivator to take part in the card exchange. Participants reported enhanced personal fulfilment, leading to improvements in wellbeing. Receiving a card could make people feel special and cherished, which was reported to establish a sense of connection with others, with potential benefits for reducing loneliness.

**Conclusions:** This study provided preliminary evidence that the KBP project might improve wellbeing, loneliness, sense of belonging and hope. Sending a kindness card in this project played a predominant role in wellbeing enhancement, and receiving a kindness card could reduce loneliness. This study suggests that the KBP project can be replicated in more contexts in the future, and might improve wellbeing and loneliness in large communities.

## 1. Introduction

This paper reports a mixed methods evaluation of an innovative participatory public health programme: the Kindness By Post (KBP) project, in which participants send and receive cards with a message of kindness from another participant. KBP aims to enhance wellbeing and social connection and reduce feelings of loneliness for those taking part.

Wellbeing typically consists of two components: 1) subjective wellbeing, which emphasises positive affective experiences, including life satisfaction, positive emotion and absence of negative mood (Ryan and Deci, 2001, p143, 144) and 2) psychological wellbeing, which relates to positive psychological functioning, comprising self-acceptance, autonomy, environmental mastery, personal growth, positive relationships and life purpose (Ryff, 1989, p1077). Wellbeing plays an essential role in quality of life. For example, a low level of wellbeing is a risk factor in developing depression (Wood and Joseph, 2010), whereas a high level of wellbeing can be a protective factor to reduce the risks for various mental illnesses and physical diseases (Ryff, 2014). Apart from health, positive wellbeing may create many necessary characteristics and resources that lead to successful outcomes in work and romantic relationships (Lyubomirsky et al., 2005a, p39). Therefore, growing evidence demonstrates that it is worthwhile enhancing wellbeing because it brings desirable benefits to people’s lives.

Loneliness is a subjective negative emotional state that arises when people perceive a discrepancy between a desired and actual social network (de Jong Giervel, 1998; Perlman and Peplau, 1981, p39; Valtorta and Hanratty, 1981, p518). Loneliness is conceptualised as multi-dimensional, consisting of intimate, relational and collective dimensions (Cacioppo et al., 2015). Consequently, loneliness refers to not only an absence of a desired interpersonal affection and intimacy with a significant love, friends and family, but also a lack of connection to a broader community. For the general population, experiencing loneliness is quite common. Approximately 10–15% of people in Europe and 20–30% of people in the UK reported experiencing loneliness frequently or most of the time (Jopling and Sserwanja, 2016). For people with worse wellbeing, feeling lonely is even more common (Kearns et al., 2015). Loneliness has negative impacts on physical health. For instance, feeling lonely weakens ongoing anabolic processes (Cacioppo and Hawkley, 2003) and has negative impacts on the immune and cardiovascular system, which increases heart disease and mortality risk (Murberg, 2004). Loneliness is also linked to increased risk of and poorer recovery from a range of mental health problems (Hawkley and Cacioppo, 2010, p219; Wang et al., 2018, p11). Loneliness may make people perceive that they have poor social skills and lead to low self-esteem (Cacioppo et al., 2000), which may consequently decrease wellbeing (Apaolaza et al., 2013, p5). Additionally, people feeling chronically lonely may become more pessimistic and encounter emotional dysphoria (Cacioppo et al., 2000), and loneliness has been found to increase the risk of later depression (Lee et al., 2021). Given the negative impacts of loneliness on physical and mental health, it is critical to prevent or reduce loneliness for individuals, which may reduce the burden on public health.

Various psychological interventions can improve wellbeing. For instance, a positive emotion focused wellbeing therapy can improve wellbeing and self-acceptance (Ruini et al., 2006). Loving-kindness meditation in which participants direct their feelings of love and compassion toward an imaginary stranger can also increase the acceptance to the self and social connectedness to others, promoting participants’ wellbeing (Hutcherson et al., 2008). Although psychological interventions can improve wellbeing with a moderate effect size (Weiss et al., 2016), most interventions require support from expert clinicians, which is expensive and might not be accessible to all people. Apart from professional interventions, people can improve wellbeing through their own activities, either achieved independently or facilitated by organised non-clinical support. The evidence based ‘Five ways to mental wellbeing’ model, promoted widely in UK helping agencies, encourages connecting with others, being physically active, learning new skills, acts of giving and kindness and mindfulness to improve wellbeing (Aked et al., 2008).

Performing kind acts has been found to improve wellbeing (Curry et al., 2018, p4; Layous et al., 2017; Layous et al., 2013, p1299; Otake et al., 2006; Kaffke, in press). Such kindness may include holding a door for another, greeting strangers or helping others with academic work (Ouwennel et al., 2014). It is suggested that kind behaviours that are courteous or altruistic may make people realise their abilities to help others, which cultivates positive feelings to themselves (Lyubomirsky et al., 2005b). Consequently, positive experiences may promote positive emotions in the long term, leading to a higher level of wellbeing (Pressman et al., 2015). As well as the person performing the kind act, people who receive kindness could also have improved wellbeing. People who received kindness have been found to show higher levels of smiling expressions, which reflected more sincere joy, compared to those who did not interact with people performing kind acts (Pressman et al., 2015). A thematic analysis found that receiving kindness could increase wellbeing beyond experiencing pleasure but also self-confidence, self-actualisation and sense of mastery (Filep et al., 2017). In addition to enhancing wellbeing, acts of kindness may also connect the giver and receiver because the receiver may feel acknowledged and valued by the giver. Furthermore, engaging in something new such as doing creative work is also encouraged as a way to improve wellbeing. Conner et al (2018) demonstrated that people who had done more creative activities (e.g., artistic ones) or developed original ideas reported a higher level of daily flourishing. Therefore, creative acts may help to achieve a positive mood and improve wellbeing, particularly if these acts also provide an opportunity to give and receive kindness.

If kind acts can enhance people’s wellbeing, it is worthwhile organising such kindness activities into more extensive and comprehensive programmes in the general population (Sin and Lyubomirsky, 2009). Previous research has only demonstrated an overall small positive effect size of kindness acts on primarily the actors (Curry et al., 2018, p19), and there is limited evidence showing the psychological impacts on receiving kindness from strangers. There is a lack of evidence regarding the effects of kindness programmes where people both perform kind acts to strangers and receive kindness from strangers, and whether this mutuality leads to bigger impacts on wellbeing and loneliness than only giving or receiving kindness. Importantly too, although there is growing evidence suggesting that kindness to strangers leads to wellbeing promotion (Dunn et al., 2008), there is still insufficient understanding about the potential mechanisms underlying the relationships between kind acts towards strangers and enhancement of people’s wellbeing. Therefore, it is essential to further investigate the experiences of kind acts for improving wellbeing. Furthermore, kindness behaviours can provide social support in which people encounter social interactions. Cacioppo et al (2015) noted that actions that provide mutual social support and increase social interactions with others could reduce loneliness. Concerning the strong associations between loneliness and wellbeing (Emerson et al., 2020; Houghton et al., 2016), it is also worth determining whether simple kindness behaviours could build connectedness between individuals, which may be an effective means to reduce loneliness.

This study will add to the developing evidence base regarding acts of kindness to and from strangers in promoting wellbeing and reducing loneliness. It aims to examine the effectiveness of a brief, self-administrated kindness programme that was organised among the general population. The public health programme ‘Kindness by Post’ (KBP) is run nationally online across the UK by the Mental Health Collective (MHC), a non-profit community interest company. In the KBP project, participants send a handmade or bought card that includes kind messages to a randomly allocated stranger, and receive a similar card from a different randomly allocated stranger, who is likely to be a different person. It has been used in a variety of social contexts, including for new students at university, for the public during the Covid-19 lockdown, and for people observing Ramadan. This study used data collected in a card exchange for Valentine’s Day in 2020—the ‘Great British Valentine’ (GBV). The exchange sought to help participants at a time which may be difficult for many, as people without a partner or in a troubled romantic relationship may experience low mood or loneliness during the Valentine’s Day period (Otnes et al., 1994). The KBP project mobilises several mechanisms for improving wellbeing, discussed above. First, it involves an act of kindness to a stranger, which has an established evidence base for improving wellbeing. Second, in contrast to most random acts of kindness projects, KBP also has a reciprocal element of giving and receiving, which may increase connections with others. Third, the creative element of card-making and kind message-writing in KBP may also be helpful for wellbeing promotion. As an inexpensive, potentially highly scalable programme, it is therefore of substantial interest to evaluate the KBP project and understand how it is experienced by participants.

Mixed methods, combining quantitative and qualitative approaches, were used to evaluate the KBP project. There are two main research questions in the quantitative analysis: (1) What are the impacts of taking part in the KBP project on participants’ wellbeing, loneliness, hope and sense of belongingness; and (2) whether baseline loneliness and the extent of participation in the card exchange relate to wellbeing changesã Our primary hypothesis is that participants would have an increase in wellbeing, measured by the Short Warwick-Edinburgh Mental Wellbeing Scale (SWEMWBS) (Stewart-Brown et al., 2009), from baseline to follow-up after taking part in the KBP project. Regarding secondary outcomes, we hypothesise that participants’ scores on measures of loneliness, hope and belongingness will improve from baseline to follow-up following the card exchange. Additionally, it is hypothesised that people with lower baseline loneliness scores would have more improvements in wellbeing at follow-up. It is also predicted that people who both sent and received a card would have more improvements in wellbeing compared to those who partially took part in the programme (who only gave or only received a card).

We will use qualitative analysis of participants’ free texts online comments to explore their experience of this programme, its perceived benefits and the potential mechanisms by which any perceived effects may have been achieved.

## 2. Materials and methods

### 2.1 Study design and setting

The current study is a secondary analysis based on the existing data collected by the KBP programme organisers in the 2020 Great British Valentine (GBV) card exchange. The research comprises a cohort study, employing a within-subject design.

### 2.2 Participants

All participants in GBV who completed pre- and post-outcome measures were included in this study. To take part in GBV, people had to be aged 16 years or above with a postal address in the UK; there were no other exclusion criteria. Participants were required to sign up for GBV online. Consistent with our ethical approval, the current study only used the data from the adult participants, aged 18 or above.

### 2.3 Ethical approvals

The study was approved by the UCL Research Ethics Committee on 9th July 2020 (REC reference 18307/001).

### 2.4 Procedure

The GBV card exchange was broadly advertised in newspapers, broadcast and social media. People who were interested in this project could sign up on the MHC website. Participants registered to take part were first invited to complete the ‘Before questionnaire’ online. They were informed that the data could be shared with external organisations anonymously for research purposes. People gave their consent to this by proceeding with the questionnaire. The baseline data collection was conducted from the 12^th^ of January to the 14^th^ of February 2020. One week before Valentine’s Day, each participant was asked to send a homemade card or letter with goodwill messages to a stranger who was randomly allocated by a computer algorithm. The stranger’s postal address and instructions regarding how to send a card were sent to the participant’s account. In return, each participant would receive a card from another stranger during the week of Valentine’s Day. If participants had not received a card, there was a back-up system that allowed participants to ask the programme organisers to arrange for a ‘replacement’ card from a volunteer. Participants were informed that there was no guarantee of receiving a card because the sending process from the stranger was completely voluntary, and not receiving a card was nothing personal. Participants could withdraw from the project at any time they wished. After the card exchange, participants were contacted again by email on the 26^th^ February and invited to complete the online ‘After Questionnaire’. They were reminded again about their anonymised data being shared and that they could give their consent by completing the questionnaire. Participants were sent a second reminder by email if they did not respond to the questionnaires. The follow-up data collection was closed on the 2^nd^ March 2020. Participants responding outside the data collection windows were excluded from the analysis.

### 2.5 Measures

At baseline and follow-up, participants completed online self-report measures of:

- Wellbeing, using the 7-item Short Warwick-Edinburgh Mental Wellbeing Scale (SWEMWBS) (Stewart-Brown et al., 2009);
- Loneliness, using the 3-item University of California at Los Angeles (UCLA) Loneliness Scale version-3 (Russell, 1996);
- Belongingness, using four items drawn from the General Belongingness Scale (GBS) (Malone et al., 2012);
- Hope, using a single item from the Beck Depression Inventory (Beck et al., 1996).

Participants reported their gender, ethnicity and age group at baseline. They reported whether or not they had sent and had received a card and provided brief free-text feedback about their experiences of the project at follow-up. There were four questions covering the specific sending or receiving experiences as well as their overall impressions and suggestions for the project. Further information regarding the study measures and how they were scored is provided in supplementary material 1.

All data were downloaded into a Microsoft Excel file by KBP staff. Free-text data were checked to remove any personally identifying information, such as names. Multiple and duplicate responses from the same person were identified by checking the sources such as email addresses of the responses. For participants who completed the measures more than once at the same timepoint in either before- or after-questionnaires, all their responses were removed, unless the responses at the same timepoint were identical, then one of the responses was saved. An anonymised dataset was thus produced, containing no personal identifiers or codes that could be used to link the data back to identifiable individuals. This anonymised dataset was then shared with the researcher at UCL through the secure UCL Dropbox system.

### 2.6 Quantitative analysis

We summarise demographic characteristics of the sample as well as baseline and follow up measures of wellbeing, loneliness, sense of belonging and hope using descriptive statistics. To explore how representative our sample was of GBV participants, we compared participants who had completed both before and after questionnaires with those who had only completed the before questionnaire using linear regression and chi squared tests.

We calculated standardised scores for wellbeing, loneliness and sense of belonging at baseline and follow up, standardised by the mean and standard deviation of the measure at baseline. For each participant, we calculated changes in wellbeing, loneliness and sense of belonging from baseline to follow up for scores on both original and standardised scales as outcomes for the analysis. Change in hope was recategorised into three groups (negative change, no change and positive change). A new binary variable was generated based on the sending and receiving experiences of the participant to represent the level of participation in the programme (full vs partial; see supplementary material 1).

We estimated change in wellbeing, loneliness and sense of belonging from baseline to follow up using separate linear regression models. Results are presented on both the original measurement scale and as standardised effect sizes. We used multinomial logistic regression to examine whether there was any improvement in hope after taking part in the programme. We assessed whether the four outcome measures are distinct from each other at baseline using Pearson correlation coefficients for the continuous measures and Spearman correlation coefficients for comparisons involving the ordinal measure of hope.

To explore whether the programme’s effectiveness was associated with either baseline loneliness or full vs partial participation, we fitted separate univariable linear regression models with change in wellbeing as the outcome and baseline loneliness and participation level respectively as the single explanatory variable. We checked the assumptions of regression models through the construction of appropriate histograms and normal quantile plots. All analyses were performed using Stata v16.

### 2.7 Qualitative analysis

The current study uses the standards for reporting qualitative research (SRQR) to report the qualitative analysis (O’Brien et al., 2014). This study used a thematic analysis to capture the pattern of the meaning of the experiences and feedback reflected by the participants. The qualitative analysis processes were guided by the thematic approach developed by Braun and Clarke (2012). There were 388 participants who completed the after-questionnaire, which contained the free-text responses, and the current analysis used the transcripts of the 289 participants who completed both before- and after-questionnaires (see Figure 1). However, the other 99 transcripts were also checked once a coding frame had been developed to determine whether there were any additional novel and distinctive codes generated. The qualitative transcripts were analysed using NVivo software version 12.

**Figure 1:**
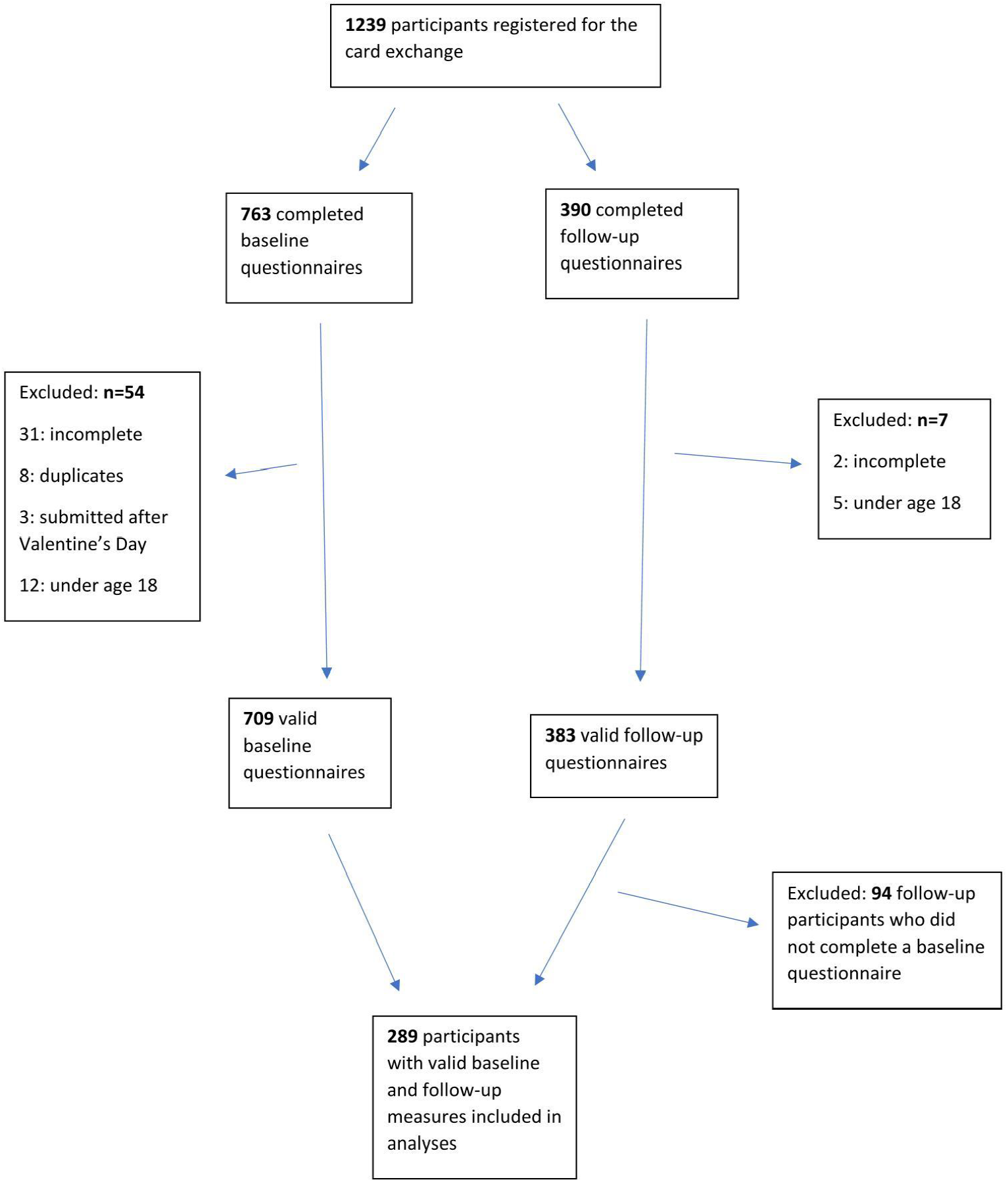
Participants included in the kindness By post analyses.

#### 2.7.1 Unit of analysis

All free-text comments to the four questions were merged as an individual transcript for each participant. All 289 participant transcripts were analysed.

#### 2.7.2 Researcher characteristics and reflexivity

The lead researcher (CW), who had the main role in coding the transcripts, has an academic psychology background. Her personal experience of loneliness and lack of belongingness during time living abroad alone for several years made her interested in determining whether the KBP project made people feel more connected to another. The other two researchers involved in analysis of qualitative data are academic researchers with anthropology (EP) and social work (BLE) backgrounds and are the Coordinator and a Co-investigator of the UKRI-funded Loneliness and Social Isolation in Mental Health Research Network, respectively. They were involved in the discussion regarding the coding framework and brought their perspectives from their own personal, professional and academic experiences to the analysis. All the researchers had participated in KBP, which helped them better understand what people reflected in the transcripts.

#### 2.7.3 Data analysis

The current study utilised both inductive and deductive approaches. Regarding the inductive analysis, the lead researcher first read through all the transcripts to become intimately familiar with the data sets’ contents and made some preliminary notes on the initial insights relevant to the research questions. For deductive analysis, there were some preliminary concepts (supplementary material 2) regarding the potential impacts of the KBP project proposed by the key stakeholder, the MHC. The preliminary concepts were considered as codes while analysing the qualitative data. Codes relevant to the research questions were generated inductively and deductively based on the semantic meaning of the responses and the latent meaning or the interpretation of the contents. The codes could be modified iteratively throughout the coding process to accommodate new ideas. After the codes were created, a cluster of codes sharing unifying features were combined into a higher-level subtheme or theme depending on how well it described a coherent and meaningful pattern in the data. The codes arising from the transcripts and the theme framework were discussed with other researchers to achieve a consensus, which would enhance the trustworthiness and credibility of the results. Themes concerning the research questions were reviewed and adjusted to capture better the overall tone of the entire dataset. Finally, the patterns and relationships of the themes were interpreted. The participation level (full or partial) was also added as an attribute of classification to the participants. Commonalities and variations of the themes were compared between participants in different participation groups.

## 3 Results

### 3.1 Quantitative results

There were 1239 participants who registered online to take part in the KBP card exchange, of which 709 had valid baseline measures, and 289 had both valid baseline and follow-up measures. Details regarding the number of individuals at each stage of the project are provided in Figure 1.

#### 3.1.1 Descriptive characteristics

For the participants in our study who completed both questionnaires, most people (N=254, 88%) were aged between 18–60 years, with equal numbers (44%) in the 19-40 and 41-60 categories. Most participants were female (N=271, 94%) and white (N=278, 96%). The mean wellbeing score for these participants was 20.7 (SD=3.48) at baseline and was 21.5 (SD=3.86) at follow-up. Eighty one percent (N=229) of participants sent and received a card, and 19% (N=54) only sent but did not receive one. Hardly anybody (N=6) received a card but did not send one. Compared to the people who only completed the before-questionnaire, participants completing both questionnaires were generally older and had a lower baseline sense of belonging. There was also weak evidence that more completers were female. There was no evidence of any other differences between completers and non-completers. Table 1 shows participants’ descriptive statistics summarised according to completer status.

**Table 1:** Demographic characteristics and baseline measures of participants.

|  |  | Non-completers: (N=420) | Completers: (N=289) | Difference between non-completers vs completers |  |
| --- | --- | --- | --- | --- | --- |
| Demographic characteristics |  |  |  |  |  |
|  |  | N(%) | N(%) | *P value |  |
| Age (years) | 19–40 | 32(55%) | 127(44%) | 0.007 |  |
|  | 41–60 | 156(37%) | 127(44%) |  |  |
|  | Over 60 | 32(8%) | 35(12%) |  |  |
| Gender | Female | 372(89%) | 271(94%) | 0.061 |  |
|  | Male | 41(10%) | 16(6%) |  |  |
|  | Non-binary, prefer not to say | 7(2%) | 2(1%) |  |  |
| Ethnicity | White | 384(91%) | 278(96%) | 0.090 |  |
|  | Black | 6(1%) | 0(0%) |  |  |
|  | Asian | 16(4%) | 5(2%) |  |  |
|  | Mixed | 10(2%) | 4(1%) |  |  |
|  | Other | 4(1%) | 2(1%) |  |  |
| Baseline measures |  |  |  |  |  |
|  |  | Mean(SD) | Mean(SD) | Estimated difference (95%CI) | #P value |
| Wellbeing |  | 20.4(3.61) | 20.7(3.48) | 0.41(-0.24 to 0.83) | 0.283 |
| Loneliness |  | 6.02 (1.87) | 5.99(1.83) | -0.63(0.30 to 0.25) | 0.848 |
| Sense of belonging |  | 18.9 (5.42) | 17.8(5.34) | -0.95(-1.92 to -0.31) | 0.007 |
| Hope |  | N(%) | N(%) | *P value |  |
|  | 0 | 17(4%) | 9(3%) | 0.726 |  |
|  | 1 | 54(13%) | 34(12%) |  |  |
|  | 2 | 129(31%) | 83(29%) |  |  |
|  | 3 | 220(52%) | 163(56%) |  |  |

**Notes:** Non-completers=Participants completing only before-questionnaire. Completers=Participants completing both before- and after-questionnaires. N=number. %=percentage. SD= standard deviation. \* Pearson Chi-squared test. # Linear regression model. Responses for hope were coded as 0, 1, 2 and 3, and greater score means higher level of hope.

#### 3.1.2 Analysis for research question 1

A simple linear regression provided strong evidence that participants had greater wellbeing after taking part in the programme (estimated change from baseline to follow-up: 0.77; 95% CI: 0.44 to 1.10; p<0.001; standardised effect size=0.21). For the secondary outcomes, the Pearson and Spearman’s rank correlation tests showed that the baseline measures of wellbeing, loneliness, sense of belonging and hope were not collinear to each other (|r|<0.7, see Table 2, Dormann et al., 2013). There was strong evidence that loneliness scores decreased from baseline to follow-up (estimated change: -0.28; 95% CI: -0.43 to -0.13; p<0.001; standardised effect size=0.15), and sense of belonging also improved (estimated change: 1.98; 95% CI:1.44 to 2.52; p<0.001; standardised effect size=0.37). There was strong evidence that the risk to have no change in hope was more than three time the risk to have positive change (relative risk ratio for no change vs positive change=3.12, 95% CI: 2.36 to 4.13, p<0.001). However, although a large majority (N=203, 70%) reported no change in hope, there was strong evidence that participants in the KBP programme were three times more likely to experience an increase in hope than a decrease (RR for negative vs positive change: 0.32; 95% CI: 0.20 to 0.53; p<0.001). Table 3 provides further details.

**Table 2:**
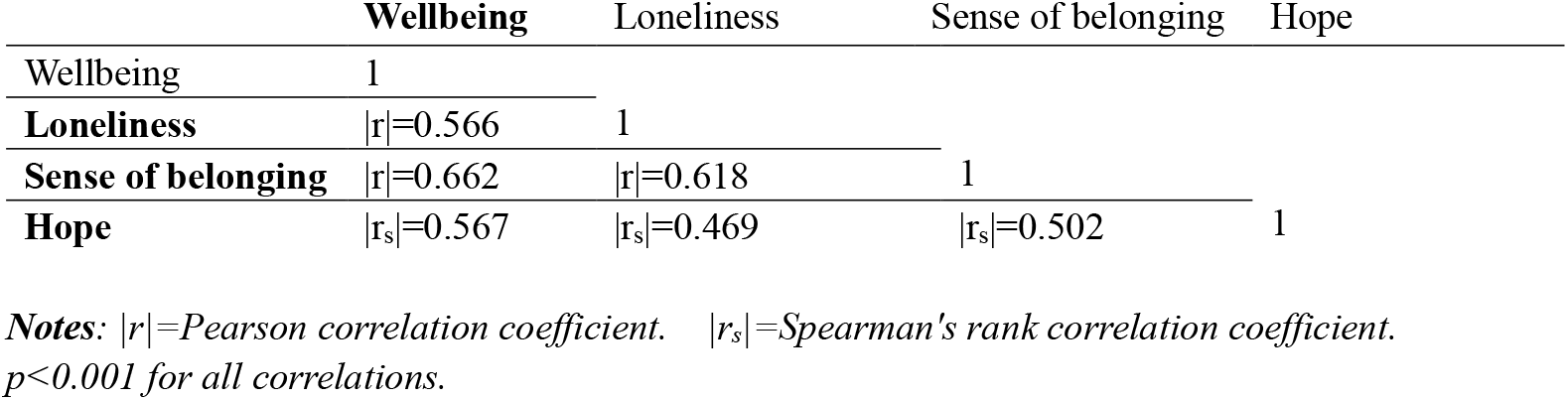
Correlations between baseline wellbeing, loneliness, sense of belonging and hope.

**Table 3:**
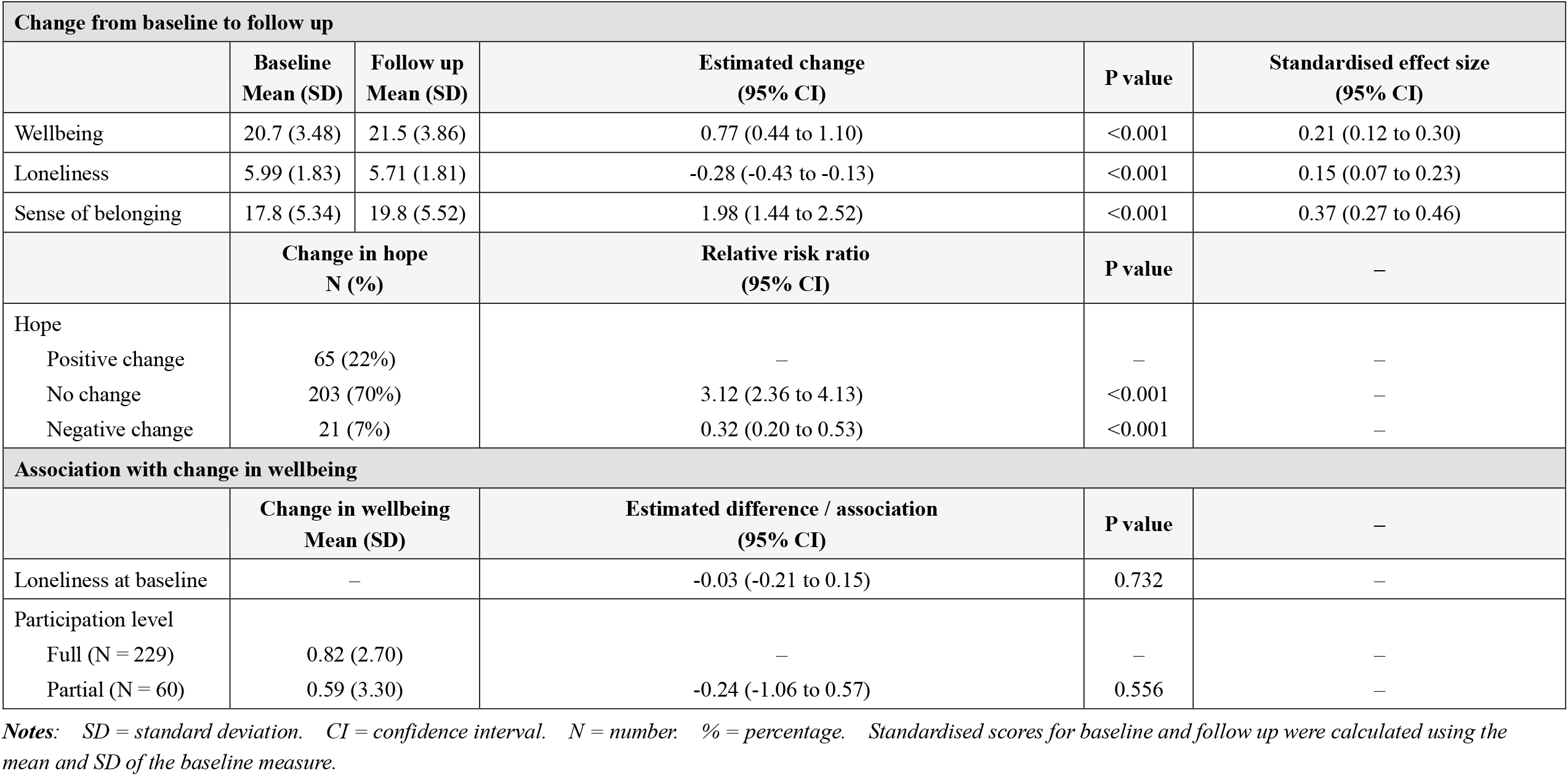
Estimated change in wellbeing, loneliness, belonging and hope and associations with change in wellbeing.

| Change from baseline to follow up |  |  |  |  |  |
| --- | --- | --- | --- | --- | --- |
|  | Baseline<br>Mean (SD) | Follow up<br>Mean (SD) | Estimated change<br>(95% CI) | P value | Standardised effect size<br>(95% CI) |
| Wellbeing | 20.7 (3.48) | 21.5 (3.86) | 0.77 (0.44 to 1.10) | <0.001 | 0.21 (0.12 to 0.30) |
| Loneliness | 5.99 (1.83) | 5.71 (1.81) | -0.28 (-0.43 to -0.13) | <0.001 | 0.15 (0.07 to 0.23) |
| Sense of belonging | 17.8 (5.34) | 19.8 (5.52) | 1.98 (1.44 to 2.52) | <0.001 | 0.37 (0.27 to 0.46) |
|  | Change in hope<br>N (%) |  | Relative risk ratio<br>(95% CI) | P value | — |
| Hope |  |  |  |  |  |
| Positive change | 65 (22%) |  | — | — | — |
| No change | 203 (70%) |  | 3.12 (2.36 to 4.13) | <0.001 | — |
| Negative change | 21 (7%) |  | 0.32 (0.20 to 0.53) | <0.001 | — |
| Association with change in wellbeing |  |  |  |  |  |
|  | Change in wellbeing<br>Mean (SD) |  | Estimated difference / association<br>(95% CI) | P value | — |
| Loneliness at baseline | — |  | -0.03 (-0.21 to 0.15) | 0.732 | — |
| Participation level |  |  |  |  |  |
| Full (N = 229) | 0.82 (2.70) |  | — | — | — |
| Partial (N = 60) | 0.59 (3.30) |  | -0.24 (-1.06 to 0.57) | 0.556 | — |

**Notes:** SD = standard deviation. CI = confidence interval. N = number. % = percentage. Standardised scores for baseline and follow up were calculated using the mean and SD of the baseline measure.

**Table 4:** Themes generated regarding how KBP was experienced by participants and how KBP achieved the benefits.

| **Overarching theme** | **Sub-theme** |
| --- | --- |
| **1. Motivators** | 1a. Altruism |
|  | 1b. Anticipate receiving |
|  | 1c. Difficult time |
| **2. Potential Mechanisms** | 2a. Pleasure in making a card |
|  | 2b. Pleasure in sending a card |
|  | 2c. Individual fulfilment |
|  | 2d. Appreciate other's thoughts and behaviours |
| **3. Project impacts** | 3a. Positive affective impacts |
|  | 3b. Feel the self is special and valued |
|  | 3c. Connection |
|  | 3d. Negative experiences |
| **4. Evaluations and suggestions for improvements** | 4a. Positive project evaluations |
|  | 4b. Unpredictable |
|  | 4c. Suggestions for improvements |

#### 3.1.3 Analysis for research question 2

There was no evidence that baseline loneliness was associated with wellbeing improvements (p=0.732) or that level of participation in the project was related to wellbeing changes (p=0.556). Please see Table 3 for further details.

### 3.2 Qualitative results

Four overarching themes were identified with the 289 transcripts: motivators, potential mechanisms, project impacts, evaluations and suggestions for improvements. There were no additional codes or themes added after checking the other 99 transcripts.

#### Theme 1. Motivators

The first theme captures the reasons why participants decided to take part in the project.

##### Sub-theme 1a. Altruism

Participants perceived that the KBP project could help others, which motivated them to initiate the kindness behaviours and take part. They considered that their kindness of sending a card would benefit others; for example, *‘It is a brilliant way to show kindness and help uplift a stranger (ID32)’*. Some participants also noted that taking part in the project gave them an opportunity to show care towards others, *‘I hoped the recipient knew someone was thinking about them (ID41)’*.

##### Sub-theme 1b. Anticipate receiving

Participants stated that taking part in this exchange programme enabled them to look forward to receiving a handmade card from a strange. Responders wrote that *‘I looked forward to receiving the card all week and checked the post more often than I normally would (ID206)’*.

##### Sub-theme 1c. Difficult time

Some participants reflected that they faced mental difficulties, stress or low mood when the project was advertised. Therefore, they hoped to take part in this positive project with an expectation of feeling more encouraged. One responder commented that *‘Valentine’s Day was a sad day for me this year (ID187)’*. Participants also felt lonely during Valentine’s Day, making them more willing to connect to the world, *‘As a single person, I guess I can feel a little left out on Valentine’s day (ID179)’*.

#### Theme 2. Potential mechanisms

Participants described four potential mechanisms that may influence their experience of the programme.

##### Sub-theme 2a. Pleasure in making a card

Many participants mentioned that they enjoyed the processes of making a card because they could slow down and spend time being creative and making artistic items. This process promoted self-care. Participants wrote that *‘I loved making the card & being creative (ID72); It made me think what would make me happy (ID164)’*.

##### Sub-theme 2b. Pleasure in sending a card

Some people stated that they enjoyed giving something that others might find helpful, *‘I sent two cards, and both individually handmade by me, and if it brightened someone’s day, then I’m delighted (ID28)’*.

##### Sub-theme 2c. Individual fulfilment

Participants obtained personal fulfilment by taking part in such a meaningful and national-wide project. The sending experiences made them feel proud of themselves, *‘Sending someone a card of good wishes made me feel useful (ID35)’*.

##### Sub-theme 2d. Appreciate other’s thoughts and behaviours

A substantial number of participants commented that it was really nice to receive a card from a stranger, and they appreciated others making beautiful handmade cards with thoughtful messages; they felt cared for by others, *‘Really appreciated the words and effort (ID84)’*.

#### Theme 3. Project impacts

This theme captures the perceived project impacts.

##### Sub-theme 3a. Positive affective impacts

Participants had positive changes in their mood by taking part in the project. They felt joyful, excited, warmed and inspired after the card-exchange, *‘It gave me a lovely warm feeling for days afterwards (ID144)’*.

##### Sub-theme 3b. Feel the self is special and valued

Receiving a card and performing a highly meaningful task that benefited others made people feel valued and special to themselves and others, *‘Receiving it made me feel very special (ID88)’*.

##### Sub-theme 3c. Connection

Participants reflected that the exchange programme provided an opportunity to connect to others despite being strangers. Hence, they felt less lonely: *‘I feel connected to my ‘senders’, even though I don’t know them (ID200); It made me feel less lonely in the world (ID12)’*. Participants also found that the project restored their faith in humanity: *‘Restored some faith in the kindness of people (ID162)’*.

##### Sub-theme 3d. Negative experiences

There were only few negative experiences compared to positive impacts reported in the responses. Some people felt sad when they did not receive a card, *‘I found it hard not receiving a card. Felt disappointed and sad (ID136)’*. Others felt disappointed getting an inappropriate card, *“upon opening I got a little disheartened as the person clearly hadn’t put as much effort in (ID54)’*. Additionally, the stress felt when attempting to make a good card was also a negative experience for some, *‘I felt quite pressured to create something worthy of sending (ID68)’*.

#### Theme 4. Evaluations and suggestions for improvements

This theme describes participants’ appraisal of the project and participants’ advice for improving it in the future.

##### Sub-theme 4a. Positive project evaluations

Participants commented that they loved the idea of the project, which was relatively simple in its procedures but was highly positive and spread kindness, *‘I love the idea of random acts of kindness (ID53); ‘Such a great movement (ID185)’*.

##### Sub-theme 4b. Unpredictable

Participants noted many uncertainties in the project. For example, they were unsure about the recipient’s responses when receiving the card, *‘Weird to not know how they were received (ID3)’*. Moreover, participants understood that there was no guarantee of receiving a card, which may be a risk for those who were vulnerable and did not receive a card, *‘It could devastate someone who is very lonely and depressed if they did not get one (ID72)’*.

##### Sub-theme 4c. Suggestions for improvements

Participants provided some suggestions for improving the project. For example, they considered that improving the project’s publicity and providing participants with confirmation that their card had been received, and that they would receive a back-up card if they requested one, could be useful. They suggested that the back-up system would raise a second expectation of receiving a card, but it might even be hurtful if the additional card was not received; thus, this back-up system should be further developed. Further details about the stranger, such as their age or more personalised information, might be helpful when making the cards.

Overall, few substantial differences were observed in participants’ programme experiences with different participation levels (sent and received/sent but not received/not sent but received). However, among the six participants who did not send but did receive a card, none of them identified the project could be an altruistic action, and this group exclusively reported the code of guilt (see supplementary material 3). Moreover, compared to the others, participants who sent but did not receive a card responded more about the disappointment of not receiving a card and had less positive affective emotions. People who sent and received a card reported more individual fulfilments compared to the other two groups. Further details about the themes and codes can be found in supplementary material 3.

## 4. Discussion

### 4.1 Main findings and interpretations

The current study evaluated a nationwide participatory public mental health project and has provided preliminary indications that the KBP project may help improve people’s wellbeing, loneliness and sense of belonging. The 0.77-point increase on the SWEMWBS measure for the KBP only has a small standardised effect size just above 0.2; however, this meets established thresholds for a meaningful, non-negligible change (Shah et al., 2018). The results also suggest that, although this project might not affect any change in hope for most people, taking part in the project is more likely to result in increased hope for the future than a loss of hope. Contrary to the hypotheses, the results provided no evidence that the level of loneliness at baseline affected the impacts of GBV on participants’ wellbeing. There was also no evidence of differences in wellbeing outcomes between people who sent and received a card and those who only gave or only received a card.

Although there was, on average, a small change in wellbeing and loneliness found in the quantitative results, the experiences shared in the qualitative results suggested that the experience of taking part in the project could be joyous and warm, which had quite large and sustained affective impacts for some. Qualitative and quantitative results both suggested that sending the kindness cards in this project could improve wellbeing, which further supported the evidence in the previous literature that performing acts of kindness promotes wellbeing and affective emotions (Curry et al., 2018, p4). Furthermore, the qualitative results indicated that the process of making and sending was highly positive for people, with engaging in a creative act and helping others both being important to many participants. These observations may help explain the quantitative finding that improvements in wellbeing were not different for those who only gave a card from those who also received one.

The qualitative results also revealed some potential mechanisms explaining how the KBP project may have helped people improve wellbeing and loneliness. First, the participants perceived the project to be an altruistic action benefiting others, which motivated them to send cards to strangers. Participants could increase personal fulfilment by thinking that they were performing a significantly meaningful task to help others, increasing their self-esteem and happiness. This finding aligns with the previous literature noting that people could derive satisfaction and gain more resources from the kindness behaviours that help others, which makes them happy (Curry et al., 2018, p11).

The findings also illustrated that enjoyment in making cards allowed people respite from the pressures of life and spend time being creative, which promoted self-care and made them feel joyful. This observation agrees with some studies proposing that engaging in creative activities may enhance positive mood and make people flourish (Dunn et al., 2008; Forgeard and Eichner, 2014). The card-making processes enabled people to search for positive quotes, poems and goodwill messages to write kind words, and it also allowed them to make an artistic card creatively. Lomas (2016) suggested that art and literature integrate the essence of humanity, and such artistic expression and appreciation helps people make sense of their lives and enriches their experiences, both of which can substantially improve wellbeing. Ryan and Deci (2000) also proposed that engaging in creative activities could satisfy the need for autonomy, which may boost wellbeing. Creatively developing good ideas offers a sense that one could master a piece of work. This self-sufficiency might evoke the positive emotions of pleasure and pride (Amabile et al., 2005, p369), which are the key components in wellbeing enhancement.

Gratitude for others’ efforts for the kind messages and handmade cards was also shown to not only make people feel excited and warmed while receiving, but it also enabled them to feel special and cherished. Consequently, participants could establish a close connection with the sender and the world because they felt cared for and loved by others. The benefits of receiving kindness that have been evidenced were mainly about the positive affective impacts (Pressman et al., 2015, Lyubomirsky and Layous, 2013; Nadler, 2017) or one’s self-efficacy, feeling of fulfilment and closeness in intimate relationships (Gleason et al., 2003, p1042). The qualitative results in this study suggested that receiving kindness from the KBP could improve wellbeing, and it could also help with the collective aspects of loneliness that people felt more included in society. However, due to the positive impacts of receiving kindness, the experiences of not receiving a card might be a potential barrier to the project’s benefits for some people.

### 4.2 Strengths and limitations

This study is novel in utilising a mixed-methods design to understand how kindness acts in a public mental health project improve wellbeing and reduce loneliness for both the giver and the receiver. It provides insight into the potential mechanisms explaining which components in the kind acts could enhance wellbeing and reduce loneliness. It also has the significant advantage of using nationwide data to explore the effectiveness of a public programme involving acts of kindness for promoting general wellbeing and social connection in a large social community.

Despite these strengths, there are still some limitations identified in this study. First, there was no control group in this pre-post study. As a result, it is not possible to draw strong inferences about the effectiveness of the KBP intervention (Fitz-Gibbon and Morris, 1987), i.e., it is unclear whether the improvements in wellbeing and loneliness were entirely attributed to the impacts of taking part in KBP, or people merely felt better after Valentine’s Day – for instance, because Valentine’s Day was over, or with the flourish of spring.

Regarding the study measures: to maintain a good response rate, the questionnaire was designed to be sufficiently brief to capture the four individual outcomes (Edwards et al., 2002). Thus, there were a limited number of questions extracted from the structured measures for belongingness and hope, which might potentially reduce the measures’ reliability (Goodman et al., 2015). This brief-measure issue was particularly prominent when analysing hope. Merely including a single item made it less sensitive for discriminating the change in hope over time, which might explain why most participants reported no change in hope. Moreover, this study only used brief online free-text responses for the qualitative analysis. The content in these materials was not always clear or in-depth; hence, it might be difficult to capture a full understanding of the experiences of the project.

Regarding the participants of KBP, participants in the current datasets were mostly white and female, potentially because the project was called ‘Great British Valentines’, which failed to attract some ethnic minority groups from distinct cultures or religions that do not celebrate Valentine’s Day. Therefore, the results might not generalise to ethnic minorities or to men. In addition, the insufficient number of people from non-White British ethnic groups does not allow us to explore whether this cultural homogeneity may enhance the programme’s effects, if a card is received from someone with some shared cultural experience and perspectives, or conversely whether exchanges with people different from oneself are even more connecting and powerful. Furthermore, we lacked data about other characteristics of interest for participants, for instance their socio-economic or marital/partnership status, with which to describe our sample or explore potential moderators of the programme’s effects. Regarding the data available to the researchers, participants who continued to complete the questionnaire at follow-up were generally older and had a lower level of belongingness than those who only completed the baseline measures. Therefore, there might be an attrition bias in the study data. Additionally, there were only six people who did not send but received a card. Therefore, this study might miss the experiences shared by this group, and whether the KBP helped them was unclear. Finally, the collection date for the ‘After questionnaire’ was only one week after the intervention. This study does not tell us whether the enhancement in wellbeing and decrease in loneliness due to this project would be maintained over the longer term.

### 4.3 Implications

#### 4.3.1 Implications for practice

The current study has shown that the KPB project has the potential to enhance wellbeing and reduce loneliness for the general population. This supports providing more KBP card exchanges in more contexts in the future, particularly during periods when people are vulnerable to mental or physical difficulties due to social isolation or natural disasters (Emerson et al., 2020), although sufficiently powered randomised controlled trials are required to provide more robust evidence on efficacy. It could be an inexpensive intervention to improve public wellbeing and reduce loneliness worldwide for people under the current social isolation orders due to the Covid-19 pandemic (Clair et al., 2021; Grover et al., 2020). However, our study suggests it may be helpful to strengthen some procedures to maximise benefits and mitigate any negative experiences of the KBP project. First, our qualitative findings suggest that not receiving a card may reduce the likelihood of project benefits. Therefore, strengthening the back-up system to provide an additional card may be helpful, to ensure that everyone could receive a card. It may also be helpful to set up a way for participants to confirm online that they have sent a card; otherwise, they could receive a reminder. Additionally, it might be helpful for participants to know whether their card was received by the recipient in a direct or indirect feedback system. People may derive satisfaction from their kindness behaviours that are appreciated by others, potentially enhancing the project’s effectiveness (Curry et al., 2018, p11; Ouweneel et al., 2014).

#### 4.3.2 Implications for research

Most importantly, it is desirable to utilise a more robust design in future evaluations, such as randomised control trials that introduce a comparison group to obtain more robust evidence of the project’s effectiveness. To explore the generalisability of our results, further research could recruit more participants with various demographic characteristics and cultural backgrounds in other KBP projects (such as currently planned MHC projects aimed at Pentecostal Christian churches over Easter, a new trial of Ramadan KBP or one for elderly people in care homes) to obtain more evidence across a broader population. Comparisons between demographic subgroups could potentially explore the influence of cultural homogeneity on the effectiveness of the KBP. It is also of interest for future evaluations to include a wider range of measures including not only wellbeing, loneliness but also fulfilment, self-esteem and the positive affective emotions that were the project’s impacts as reported in the current qualitative results. More in-depth qualitative interviews with participants are also necessary to help understand the mechanisms and experiences better.

Another intriguing direction for future research would be to analyse how long the positive outcomes are maintained after the kindness interventions. The creative processes in the kindness acts that promote wellbeing and affective emotions might last no more than two days (Conner et al., 2018; Amabile et al., 2005). Ouweneel and colleagues (2014) suggested that the effects of kindness acts could lessen as time progresses, whereas Seligman et al. (2005) argued that such effects could last for several months. Therefore, future research could potentially involve a longitudinal study tracking the KBP project’s impacts. Additionally, the positive outcomes of this project were achieved with participants whose average wellbeing scores were nearly three quarters of a standard deviation below the population norm (Ng Fat et al., 2017). Therefore, future studies could investigate whether the KBP might also work well specifically for people with depression. Finally, researchers could also perform a cost-effectiveness study for the KBP project to determine whether the modest gains in wellbeing and loneliness found in this study represent good value for money.

## 5. Conclusion

This study showed preliminary evidence that the KBP project may enhance wellbeing and reduce loneliness. The sending process seems to play a crucial role in the main positive impacts of the project. Qualitive reports suggests that altruism motivates people to initiate kindness behaviours, through which people may obtain personal fulfilment, and this could potentially enhance wellbeing. Moreover, receiving kindness enhances self-esteem and enables participants to perceive a connection with the sender who provides kindness, even though they are a stranger and there is no ongoing contact. Thus, the social connection might reduce participants’ loneliness. Therefore, this study supports providing future KBP projects, as a new initiative that is not only simple and cheap but may also be powerful for wellbeing promotion and loneliness prevention in the community.

## Supporting information

KBP_Supplemental Material

## Data Availability

All data produced in the present study are available upon reasonable request to the authors

## List of abbreviations

KBP: Kindness by Post
GBV: Great British Valentines
SWEMWBS: Short Warwick-Edinburgh Mental Wellbeing Scale
UCLA: University of California at Los Angeles
GBS: General Belongingness Scale
UCL: University College London

## Consent for publication

Not applicable

## Ethics approval and consent to participate

The study received ethical approval from UCL [reference number: 18307 001]. Participation in the study was voluntary and all participants whose data was analysed consented by proceeding the questionnaires to take part.

## Availability of data and materials

The datasets generated and/or analysed during the current study are not publicly available due the protection of the confidentiality of research participants, but are available from the corresponding author on reasonable request.

## Authors’ contributions

CW performed the data analysis, drafted the ethical application forms and the research report. BL and EP helped proceed the ethical approval, revised the qualitative coding framework and provided feedback to the report to improve the writing-up. RJ helped with the quantitative data analysis. All authors read and approved the final manuscript.

## Acknowledgements

We thank Dr Amy Pollard, the Director of the Mental Health Collective and organiser of the ‘Kindness by Post’ project, for her support for this study, for providing access to the study data and for contributing to development of themes for the qualitative analysis.

We also extend thanks to the participants for contributing their data to the current research.

## References

Aked, J., Marks, N., Cordon, C. & Thompson, S. (2008). Five Ways to Wellbeing: A report presented to the Foresight Project on communicating the evidence base for improving people’s well-being. London: New Economics Foundation.

Amabile, T. M., Barsade, S. G., Mueller, J. S., and Staw, B. M. (2005). Affect and creativity at work. Administrative science quarterly, 50(3), 367–403. doi:10.2189/asqu.2005.50.3.367

Apaolaza, V., Hartmann, P., Medina, E., Barrutia, J.M., and Echebarria, C. (2013). The relationship between socializing on the Spanish online networking site Tuenti and teenagers’ subjective wellbeing: The roles of self-esteem and loneliness. Computers in Human Behavior 29(4), 1282–1289. doi: 10.1016/j.chb.2013.01.002

Baumeister, R. F., and Leary, M. R. (1995). The Need to Belong: Desire for Interpersonal Attachments as a Fundamental Human Motivation. Psychological Bulletin 117, 497–529. doi:10.1037/0033-2909.117.3.497.

Beck, A.T., Steer, R.A., and Brown, G.K. (1996). Beck depression inventory (BDI-II). Pearson.

Braun, V., and Clarke, V. (2012). Thematic analysis. APA handbook of research methods in psychology, Vol 2: Research designs: Quantitative, qualitative, neuropsychological, and biological., 57–71. doi:10.1037/13620-004.

Cacioppo, J. T., Ernst, J. M., Burleson, M. H., McClintock, M. K., Malarkey, W. B., Hawkley, L. C., et al. (2000). Lonely traits and concomitant physiological processes: the MacArthur social neuroscience studies. International journal of psychophysiology : official journal of the International Organization of Psychophysiology 35, 143–154. doi:10.1016/S0167-8760(99)00049-5.

Cacioppo, J.T., and Hawkley, L.C. (2003). Social isolation and health, with an emphasis on underlying mechanisms. Perspect Biol Med 46(3 Suppl), S39–52.

Cacioppo, S., Grippo, A.J., London, S., Goossens, L., and Cacioppo, J.T. (2015). Loneliness: clinical import and interventions. Perspect Psychol Sci 10(2), 238–249. doi: 10.1177/1745691615570616.

Clair, R., Gordon, M., Kroon, M., and Reilly, C. (2021). The effects of social isolation on well-being and life satisfaction during pandemic. Humanities and Social Sciences Communications 2021 8:1 8, 1–6. doi:10.1057/s41599-021-00710-3.

Conner, T. S., DeYoung, C. G., and Silvia, P. J. (2018). Everyday creative activity as a path to flourishing. Journal of Positive Psychology 13, 181–189. doi:10.1080/17439760.2016.1257049.

Curry, O. S., Rowland, L. A., Van Lissa, C. J., Zlotowitz, S., McAlaney, J., & Whitehouse, H. (2018). Happy to helpã A systematic review and meta-analysis of the effects of performing acts of kindness on the well-being of the actor. Journal of Experimental Social Psychology, 76, 320–329. doi:10.1016/j.jesp.2018.02.014.

de Jong-Gierveld, J. (1998). A review of loneliness: Concepts and definitions, determinants and consequences. Reviews in Clinical Gerontology, (8), 73–80. doi:10.1017/S0959259898008090.

Dormann, C. F., Elith, J., Bacher, S., Buchmann, C., Carl, G., Carré, G., et al. (2013). Collinearity: a review of methods to deal with it and a simulation study evaluating their performance. Ecography 36, 27–46. doi:10.1111/J.1600-0587.2012.07348.X.

Dunn, E.W., Aknin, L.B., and Norton, M.I. (2008). Spending money on others promotes happiness. Science 319(5870), 1687–1688. doi:10.1126/science.1150952.

Edwards, P., Roberts, I., Clarke, M., DiGuiseppi, C., Pratap, S., Wentz, R., and Kwan, I. (2002). Increasing response rates to postal questionnaires: systematic review. BMJ (Clinical research ed.), 324(7347), 1183. doi:10.1136/bmj.324.7347.1183.

Emerson, E., Fortune, N., Aitken, Z., Hatton, C., Stancliffe, R., and Llewellyn, G. (2020). The wellbeing of working-age adults with and without disability in the UK: Associations with age, gender, ethnicity, partnership status, educational attainment and employment status. Disabil Health J 13(3), 100889. doi: 10.1016/j.dhjo.2020.100889.

Filep, S., Macnaughton, J., Glover, T., Filep, S., Macnaughton, J., and Glover, T. (2017). Tourism and gratitude: Valuing acts of kindness. Annals of Tourism Research 66, 26–36. doi:10.1016/J.ANNALS.2017.05.015.

Fitz-Gibbon, C. T. and Morris L. L. (1978). How To Design a Program Evaluation. Program Evaluation Kit, 3.

Forgeard, M. J. C., and Eichner, K. v. (2014). Creativity as a Target and Tool for Positive Interventions. The Wiley Blackwell Handbook of Positive Psychological Interventions, 135–154. doi:10.1002/9781118315927.CH7.

Gleason, M. E., Iida, M., Bolger, N., and Shrout, P. E. (2003). Daily supportive equity in close relationships. Personality & social psychology bulletin, 29(8), 1036–1045. doi:10.1177/0146167203253473.

Goodman, A., Wrigley, J., Silversides, K., and Venus-Balgobin, N. (2015). Measuring your impact on loneliness in later life. London UK: Campaign to End Loneliness.

Grover, S., Sahoo, S., Mehra, A., Avasthi, A., Tripathi, A., Subramanyan, et al. (2020). Psychological impact of COVID-19 lockdown: An online survey from India. Indian journal of psychiatry, 62(4), 354–362. doi:10.4103/psychiatry.IndianJPsychiatry_427_20

Hawkley, L. C., and Cacioppo, J. T. (2010). Loneliness matters: a theoretical and empirical review of consequences and mechanisms. Annals of behavioral medicine : a publication of the Society of Behavioral Medicine, 40(2), 218–227. doi:10.1007/s12160-010-9210-8.

Houghton, S., Hattie, J., Carroll, A., Wood, L., and Baffour, B. (2016). It Hurts To Be Lonely! Loneliness and Positive Mental Wellbeing in Australian Rural and Urban Adolescents. Journal of Psychologists and Counsellors in Schools 26, 52–67. doi:10.1017/JGC.2016.1.

Hutcherson, C.A., Seppala, E.M., and Gross, J.J. (2008). Loving-kindness meditation increases social connectedness. Emotion 8(5), 720–724. doi: 10.1037/a0013237.

Jopling, K., & Sserwanja, I. (2016). Loneliness across the life course: a rapid review of the evidence. Calouste Gulbenkian Foundation, UK Branch.

Kaffke, L. (2018). Can “Acts of Kindness “enhance mental well-being and flourishing in individuals from the general Dutch population?, University of Twente.

Kearns, A., Whitley, E., Tannahill, C., and Ellaway, A. (2015). Loneliness, social relations and health and well-being in deprived communities. Psychology, health & medicine 20, 332–344. doi:10.1080/13548506.2014.940354.

Layous, K., Lee, H., Choi, I., and Lyubomirsky, S. (2013). Culture matters when designing a successful happiness-increasing activity: A comparison of the United States and South Korea. Journal of Cross-Cultural Psychology 44(8), 1294–1303. doi:10.1177/0022022113487591.

Layous, K., Nelson, S.K., Kurtz, J.L., and Lyubomirsky, S. (2017). What triggers prosocial effortã A positive feedback loop between positive activities, kindness, and well-being. The Journal of Positive Psychology 12(4), 385–398. doi:10.1080/17439760.2016.1198924.

Lee, S. L., Pearce, E., Ajnakina, O., Johnson, S., Lewis, G., Mann, F., et al. (2021). The association between loneliness and depressive symptoms among adults aged 50 years and older: a 12-year population-based cohort study. The Lancet Psychiatry 8, 48–57. doi:10.1016/S2215-0366(20)30383-7.

Lomas, T. (2016). Positive art: Artistic expression and appreciation as an exemplary vehicle for flourishing. Review of General Psychology, 20(2), 171–182. doi:10.1037/gpr0000073.

Lyubomirsky, S., and Layous, K. (2013). How do simple positive activities increase well-beingã. Current directions in psychological science, 22(1), 57–62. doi: 10.1177/0963721412469809.

Lyubomirsky, S., King, L., and Diener, E. (2005a). The benefits of frequent positive affect: does happiness lead to success? Psychol Bull 131(6), 803–855. doi: 10.1037/0033-2909.131.6.803.

Lyubomksky, S., Sheldon, K. M., and Schkade, D. (2005b). Pursuing happiness: The architecture of sustainable change. Review of General Psychology 9, 111–131. doi:10.1037/1089-2680.9.2.111.

Malone, G.P., Pillow, D.R., and Osman, A. (2012). The general belongingness scale (GBS): Assessing achieved belongingness. Personality and individual differences 52(3), 311–316. doi: 10.1016/j.paid.2011.10.027.

Murberg, T.A. (2004). Long-term effect of social relationships on mortality in patients with congestive heart failure. Int J Psychiatry Med 34(3), 207–217. doi: 10.2190/gkj2-p8bd-v59x-mjnq.

Nadler, A. (2017). The human essence in helping relations: Belongingness, independence, and status. The Oxford handbook of the human essence, 1, 123–134.

Ng Fat, L., Scholes, S., Boniface, S., Mindell, J., and Stewart-Brown, S. (2017). Evaluating and establishing national norms for mental wellbeing using the short Warwick-Edinburgh Mental Well-being Scale (SWEMWBS): findings from the Health Survey for England. Quality of life research : an international journal of quality of life aspects of treatment, care and rehabilitation, 26(5), 1129–1144. doi:10.1007/s11136-016-1454-8.

O’Brien, B.C., Harris, I.B., Beckman, T.J., Reed, D.A., and Cook, D.A. (2014). Standards for reporting qualitative research: a synthesis of recommendations. Acad Med 89(9), 1245–1251. doi: 10.1097/acm.0000000000000388.

Otake, K., Shimai, S., Tanaka-Matsumi, J., Otsui, K., and Fredrickson, B.L. (2006). HAPPY PEOPLE BECOME HAPPIER THROUGH KINDNESS: A COUNTING KINDNESSES INTERVENTION. J Happiness Stud 7(3), 361–375. doi: 10.1007/s10902-005-3650-z.

Otnes, C., Ruth, J.A., and Milbourne, C.C. (1994). The pleasure and pain of being close: Men’s mixed feelings about participation in Valentine’s Day gift exchange. ACR North American Advances.

Ouweneel, E., Le Blanc, P.M., and Schaufeli, W.B. (2014). On being grateful and kind: results of two randomized controlled trials on study-related emotions and academic engagement. J Psychol 148(1), 37–60. doi: 10.1080/00223980.2012.742854.

Perlman, D., & Peplau, L. A. (1981). “Personal Relationships: 3. Relationships in Disorder,” in the Toward a Social Psychology of Loneliness, ed. R. Gilmour, & S. Duck (London: Academic Press), 31–56.

Pressman, S.D., Kraft, T.L., and Cross, M.P. (2015). It’s good to do good and receive good: The impact of a ‘pay it forward’style kindness intervention on giver and receiver well-being. The Journal of Positive Psychology 10(4), 293–302. doi: 10.1080/17439760.2014.965269.

Ruini, C., Belaise, C., Brombin, C., Caffo, E., and Fava, G.A. (2006). Well-being therapy in school settings: a pilot study. Psychother Psychosom 75(6), 331–336. doi: 10.1159/000095438.

Russell, D.W. (1996). UCLA Loneliness Scale (Version 3): reliability, validity, and factor structure. J Pers Assess 66(1), 20–40. doi: 10.1207/s15327752jpa6601_2.

Ryan, R. M., and Deci, E. L. (2000). Self-determination theory and the facilitation of intrinsic motivation, social development, and well-being. The American psychologist, 55(1), 68–78. doi: 0.1037//0003-066x.55.1.68.

Ryan, R.M., and Deci, E.L. (2001). On happiness and human potentials: a review of research on hedonic and eudaimonic well-being. Annu Rev Psychol 52, 141–166. doi: 10.1146/annurev.psych.52.1.141.

Ryff, C. D. (1989). Happiness is everything, or is itã Explorations on the meaning of psychological well-being. Journal of Personality and Social Psychology, 57(6), 1069–1081. doi:0.1037/0022-3514.57.6.1069.

Ryff, C.D. (2014). Psychological well-being revisited: advances in the science and practice of eudaimonia. Psychother Psychosom 83(1), 10–28. doi: 10.1159/000353263.

Seligman, M. E., Steen, T. A., Park, N., and Peterson, C. (2005). Positive psychology progress: empirical validation of interventions. The American psychologist, 60(5), 410–421. doi: 10.1037/0003-066X.60.5.410.

Shah, N., Cader, M., Andrews, W.P., Wijesekera, D., and Stewart-Brown, S.L. (2018). Responsiveness of the Short Warwick Edinburgh Mental Well-Being Scale (SWEMWBS): evaluation a clinical sample. Health Qual Life Outcomes 16(1), 239. doi: 10.1186/s12955-018-1060-2.

Sin, N.L., and Lyubomirsky, S. (2009). Enhancing well-being and alleviating depressive symptoms with positive psychology interventions: a practice-friendly meta-analysis. J Clin Psychol 65(5), 467–487. doi: 10.1002/jclp.20593.

Stewart-Brown, S., Tennant, A., Tennant, R., Platt, S., Parkinson, J., and Weich, S. (2009). Internal construct validity of the Warwick-Edinburgh Mental Well-being Scale (WEMWBS): a Rasch analysis using data from the Scottish Health Education Population Survey. Health Qual Life Outcomes 7, 15. doi:10.1186/1477-7525-7-15.

Valtorta, N., and Hanratty, B. (2012). Loneliness, isolation and the health of older adults: do we need a new research agenda? J R Soc Med 105(12), 518–522. doi:10.1258/jrsm.2012.120128.

Wang, J., Mann, F., Lloyd-Evans, B., Ma, R., and Johnson, S. (2018). Associations between loneliness and perceived social support and outcomes of mental health problems: a systematic review. BMC psychiatry, 18(1), 156. doi:10.1186/s12888-018-1736-5.

Weiss, L. A., Westerhof, G. J., and Bohlmeijer, E. T. (2016). Can We Increase Psychological Well-Beingã The Effects of Interventions on Psychological Well-Being: A Meta-Analysis of Randomized Controlled Trials. PloS one 11. doi:10.1371/JOURNAL.PONE.0158092.

Wood, A.M., and Joseph, S. (2010). The absence of positive psychological (eudemonic) well-being as a risk factor for depression: a ten year cohort study. J Affect Disord 122(3), 213–217. doi: 10.1016/j.jad.2009.06.032.

