## Supplementary material for "‘Kindness By Post’: A Mixed-Methods Evaluation of A Participatory Public Mental Health Project": KBP_Supplemental Material

**Supplementary material 1: Details about the variable and their measures and the free-text questions**

| | | | | |
|---|---|---|---|---|
| **Demographic Characteristics** | | | | |
| Age | Categorical | How old are you? | Under 18/ 19-40/ 41-60/ Over 60 | |
| Gender | Categorical | What is your gender? | Female/ Male/ Non-binary or prefer not to say | |
| Ethnicity | Categorical | Which of the following best describe your ethnicity | White/ Black/ Asian/ Mixed/ Other* | |
| **Measures of wellbeing, hope, loneliness and sense of belonging** | | | | |
| Wellbeing | Continuous | 7-item Short Warwick-Edinburgh Mental Wellbeing Scale (SWEMWS):<br>Likert-scale from 1 (none of the time) to 5 (all of the time). The total scores are by first summing the scores for each of the seven items and then transformed into metric scores using the SWEMWBS conversion table. | The range is 7-35, and greater score indicates a higher positive mental wellbeing. | • High internal consistency in the UK general population (Fat et al., 2017).<br>• High one-week test-retest reliability among university students (Tennant et al., 2007).<br>• Good convergent validity. (Fat et al., 2017).<br>• Good construct and concurrent validity in Danish adults (Koushede et al., 2019).<br>• Good discriminant validity (Fat et al., 2017). |
| Loneliness | Continuous | 3 questions from UCLA loneliness scale:<br>Likert scale from 1 (hardly ever or never) to 3 (often). | Ranging from 3 to 9, with greater score indicating being lonelier. | High reliability (internal consistency and one-year test-retest reliability), good convergent validity and good |

| | | | | |
|---|---|---|---|---|
| | | | | construct validity of the UCLA loneliness measures (Russell, 1996). |
| Belongingness | Continuous | 4 questions from the General Belongingness Scale (GBS): 7-point Likert scale with one item reversed. | Ranging from 4 to 28. Higher score indicates higher level of belongingness. | High reliability and high validity of the complete GBS measure (Malone et al., 2012). |
| Hope | Categorical | A single item hope question taken from the Beck Depression Inventory with 4 responses. | Four responses are coded as 0, 1, 2 and 3. Greater score means more hope. | N/A |
| **Other variables** | | | | |
| Sending a card | Binary | Yes or no questions asking whether the participants gave a card. | Yes/ No | |
| Receiving a card | Binary | Yes or no questions asking whether the participants received cards. | Originally designed with the responses: Yes/ Received two cards/ No<br>Adjusted to Yes/ No | |
| Participation level in the programme | Binary | Whether participants fully or partially participated in the project. | Participants who sent and received the cards are grouped as full participation. The others who did not send or did not receive a card or neither received nor sent a card are grouped as partial participation. | |

Note: * The broad ethnicity categories were derived by collapsing the 18 census ethnicity categories. The ethnic groups were described as White (White/White Irish/White Gypsy-Traveller/White other), Black (Black African/Black Caribbean/Black other), Asian (Bangladeshi/Chinese/Indian/Pakistani/Asian other), Mixed (Mixed White-Asian/Mixed White-Black African/Mixed White-Black Caribbean/Mixed other) and Other.

References:

Koushede, V., Lasgaard, M., Hinrichsen, C., Meilstrup, C., Nielsen, L., Rayce, et al. (2019). Measuring mental well-being in Denmark: Validation of the original and short version of the Warwick-Edinburgh mental well-being scale (WEMWBS and SWEMWBS) and cross-cultural comparison across four European settings. *Psychiatry research*, 271, 502–509. doi:10.1016/j.psychres.2018.12.003.

Malone, G.P., Pillow, D.R., and Osman, A. (2012). The general belongingness scale (GBS): Assessing achieved belongingness. *Personality and individual differences* 52(3), 311-316. doi: 10.1016/j.paid.2011.10.027.

Ng Fat, L., Scholes, S., Boniface, S., Mindell, J., and Stewart-Brown, S. (2017). Evaluating and establishing national norms for mental wellbeing using the short Warwick-Edinburgh Mental Well-being Scale (SWEMWBS): findings from the Health Survey for England. *Quality of life research : an international journal of quality of life aspects of treatment, care and rehabilitation*, 26(5), 1129–1144. doi:10.1007/s11136-016-1454-8.

Russell, D.W. (1996). UCLA Loneliness Scale (Version 3): reliability, validity, and factor structure. *J Pers Assess 66*(1), 20-40. doi: 10.1207/s15327752jpa6601_2.

Tennant, R., Hiller, L., Fishwick, R., Platt, S., Joseph, S., Weich, et al. (2007). The Warwick-Edinburgh Mental Well-being Scale (WEMWBS): development and UK validation. *Health and quality of life outcomes*, 5, 63. doi:10.1186/1477-7525-5-63.

### Supplementary material 2: Preliminary themes generated by the Mental Health Collective for deductive approach in the qualitative analysis

| **Preliminary themes** | **Explanations** |
|---|---|
| Agency and empowerment | A sense of self-efficacy in being able to help someone else |
| Self-esteem | A positive feeling because of helping others |
| Being moved | Feeling of tearful or emotional after receiving a card |
| Inclusion | Feeling connected or accepted by an inner group |
| More positive views of the broader world | A shift in generalised perceptions, saying things like “restored my faith in humanity”; “it felt like the world was a bit softer”; “it showed me there are good people out there” |
| Destiny | Feeling a right time to receive a kindness message |

The Director of the Mental Health Collective provided help with the generation of the preliminary themes. She used to be a researcher in social anthropology but also has lived experience of receiving postal messages of kindness during a period of mental distress, which prompted her to develop the KBP project.

**Supplementary material 3: Themes and codes generated from the qualitative analysis**

| **Overarching themes** | **Sub-themes** | **Codes** | **Examples of quotes** |
|---|---|---|---|
| 1. Motivators | 1a. Altruism | Benefit others | *'It is a brilliant way to show kindness and help uplift a stranger (ID32)'.* |
| | | Care about others | *'Wanted to offer kind words of wisdom and make the person feel good about their self felt (ID120)'.* |
| | 1b. Anticipate receiving | Anticipate receiving | *'I looked forward to receiving the card all week and checked the post more often than I normally would (ID206)'.* |
| | 1c. Difficult time | Low mood | *'Came on a day I was feeling quite down (ID7)'.* |
| | | Stressed out | *'I am usually just a worker at work and a mum the whole time, and though my partner is kind we are stressed and busy all the time. In my job and role as a parent and partner I care for others and feel depleted all the time (ID46)'.* |
| | | Mental difficulties | *'I was having a bad mental health day (ID208)'.* |
| 2. Mechanisms | 2a. Pleasure in making a card | Enjoy making | *'I enjoyed designing my card which was very simple. I had a look online for suitable texts and found some really thought provoking ones (ID142)'.* |
| | | Self-care | *'It made me stop and think about what I would want someone to say to me (ID85)'.* |
| | 2b. Pleasure in sending a card | Enjoy sending | *'I sent 2 cards, and both individually handmade by me, and if it brightened someone's day, then I'm delighted (ID28)'.* |
| | 2c. Individual fulfilment | Fulfilling | *'Sending someone a card of good wishes made me feel useful (ID35)'.* |
| | | Feel good about self | *'Feeling good about ourselves for making the cards (ID48)'.* |
| | | Appreciation | *'I appreciated that someone had made an effort (ID2)'.* |

| | | | |
|---|---|---|---|
| | 2d. Appreciate other's thoughts and behaviours | Being cared | *'As though some one 'out there cared (ID143)'.* |
| | | Love the cards | *'I absolutely loved it. The words were somehow very relevant even though they were from a stranger (ID187)'.* |
| 3. Project impacts | 3a. Positive affective impacts | Warm | *'Completely positive and heart warming (ID56)'.* |
| | | Excitement | *'I felt really excited when I saw the card cone through the letterbox (ID42)'.* |
| | | Lighten one's mood | *'I still look at it and feel joy. Even typing about it is making me smile (ID140)'.* |
| | 3b. Feel the self is special and valued | Special and valued | *'Receiving it made me feel very special (ID88)'.* |
| | 3c. Connection | Less lonely | *'It made me feel less lonely in the world (ID12)'.* |
| | | Connected | *'I feel connected to my 'senders', even though I don't know them (ID200)'.* |
| | | Share humanity | *'It was a nice thing to receive and made me have a little more hope for humanity in general (ID188)'.* |
| | 3d. Negative experiences | Disappointed not receiving | *'I found it hard not receiving a card. Felt disappointed and sad (ID136)'.* |
| | | Disappointed with the card | *'The upon opening I got a little disheartened as the person clearly hadn't put as much effort in (ID54)'.* |
| | | Guilt | *'I felt so embarrassed I had forgotten all about doing this (ID81)'.* |
| | | Stress in making | *'Was a bit nervous especially about creating my own card (ID79)'.* |
| | 4a. Positive project evaluations | Great idea and concept | *'Love the idea of the whole project (ID163)'.* |

| | | | |
|---|---|---|---|
| 4. Evaluations and suggestions for improvements | | Spread kindness | *'Pleased to be involved with spreading a little kindness (ID74)'.* |
| | | Positive project | *'I feel that it was a positive experience overall & think that the project is a good one (ID122)'.* |
| | 4b. Unpredictable | Unsure about receiving | *'Weird to not know how they were received (ID3)'.* |
| | | Risk to vulnerable people | *'Because I guess if you are already feeling unloved that might make you feel even more unlovable (ID53)'.* |
| | 4c. Suggestions for improvements | Confirmatio n | *'People being able to log into their account online and click to confirm when they've created and sent a card (could give you an idea of participation ahead of the day, and a heads up for the back-up system demand)? (ID9)'.* |
| | | More publicity | *'Needs more publicity (ID96)'.* |
| | | More details about the receiver | *'The only problem I encountered was not knowing anything about my recipient...I could have made the card more personal.(ID165)'.* |
